# Diagnosis of SARS-CoV-2 in children: accuracy of nasopharyngeal swab compared to nasopharyngeal aspirate

**DOI:** 10.1101/2020.08.20.20178012

**Authors:** Giada Maria Di Pietro, Ester Capecchi, Ester Luconi, Giovanna Lunghi, Samantha Bosis, Giuseppe Bertolozzi, Barbara Cantoni, Giuseppe Marano, Patrizia Boracchi, Elia Biganzoli, Silvana Castaldi, Paola Marchisio, On behalf of Testing Pediatric Covid-19 (TPC-19), Andrea Gori, Carlo Agostoni, Raffaella Pinzani, Ferruccio Ceriotti

## Abstract

The tests currently used for the direct identification of SARS-CoV-2 include specimens taken from upper and lower respiratory tract; recommendations from Word Health Organization prioritise nasopharyngeal swab (NS). In literature there are not available paediatric studies about the identification of SARS-CoV-2 through nasopharyngeal aspirate (NPA), but the use of NPA is deemed to be better than NS to identify respiratory viruses in children. The aim of our study is to evaluate diagnostic performances of NS compared to NPA for the detection of SARS-CoV-2 in children. We collected 300 paired samples (NS and NPA) from children hospitalized and followed up in our paediatric unit. We calculated the sensitivity and specificity of NS referred to NPA of the whole sample and then, considering both the age (≥ and < 6 years old) and the period of collection (March vs follow up) as covariates in different analysis. The NS has a low sensitivity in detecting SARS-CoV-2 in children when referred to NPA; whereas its specificity results high. In children under 6 years of age, our results suggest to prefer the collection of NS, whenever possible. Though statistically not significant, the sensitivity of NS becomes higher if it is performed before NPA.

## INTRODUCTION

In December 2019 appeared in China a novel coronavirus, designated as severe acute respiratory syndrome coronavirus 2 (SARS-CoV-2), responsible for an epidemic of respiratory disease, known as Coronavirus Disease (COVID-19), which turn into an international health emergency, as declared by the World Health Organization (WHO) (https://www.who.int/docs/default-source/coronaviruse/situation-reports/20200311-sitrep-51-covid-19.pdf?sfvrsn=1ba62e57_10). In February 2020 the outbreak involved also Italy, which became one of the most affected countries by the global COVID-19 pandemic (https://www.epicentro.iss.it/coronavirus/sars-cov-2-dashboard).

Most cases of patients reported in literature concern adults, with an aging-dependent mortality rate which is higher in elderly or in subjects with chronic underlying diseases.^1^

Data regarding infected children are so far limited. Wu et al in a review about 44,672 laboratory-confirmed cases of COVID-19, showed that 2% of cases occurred in children from 10 to 19 years of age and only 1% under 9 years of age, with no deaths reported in the latter group.^2^ Furthermore, children younger than 18 years of age appeared to be less vulnerable to the infection, to have milder symptoms and less severe disease compared with adults.^2-5^ A Chinese observational study reported that, of the 1,391 children younger than 16 years of age tested for SARS-CoV-2, only 171 resulted positive; among them 15.8% were asymptomatic and the remaining part appeared to have a mild clinical course.^6^ These observations compare with recent results from the seroprevalence study in the first lockdown red area of Lombardy, showing a linear increase of the log odds IgG positivity with age, ranging from 9.1% at 5 years to 12.5% at 20 years ending around 40% for people with age over 80 years.^7^

The tests currently used for the direct identification of SARS-CoV-2 include specimens taken from the upper (nasopharyngeal/oropharyngeal swab and nasopharyngeal aspirate) and the lower respiratory tract (bronchoalveolar lavage, tracheal aspirate, sputum).^8-10^

Upper respiratory specimens are easy and less invasive to obtain, moreover their collection exposes healthcare workers to a lower risk of infection; so, they are recommended for asymptomatic children or for patients with mild symptoms. The collection of lower respiratory specimens is reserved to symptomatic or severe cases due to the discomfort caused to the infants, the special devices and skilled operators required to obtain them.^8^

Yang et al demonstrated that the identification of SARS-CoV-2 in the bronchoalveolar lavage fluid (BALF) had the 100% of positive rate when collected from severe cases, while the sputum had the highest positive rate in both severe and mild cases, followed by nasal swab.^8-12^ As regards samples collected from the upper airways, higher viral loads were detected in the nose than in the throat; indeed, recommendations from WHO prioritize nasopharyngeal swab over oropharyngeal swab.^8,13^

Zou et al demonstrated that the viral load in symptomatic children is similar to that in asymptomatic patients, suggesting the potential contagiousness of the latter. Furthermore, they detected higher viral loads in specimens collected soon after symptom onset, finding that the risk of transmission is higher in the earlier stage of infection. ^8,13^

Even if a positive test is highly suggestive of true COVID-19, a negative test does not rule out the disease.^14^ Several factors may contribute to false-negative results including the sampling technique, transportation process, limited genes detection on specimens but also the nature of coronavirus itself.^15^ Testing of specimens from multiple sites may reduce false-negative results.^11^

Currently, in literature there are not available paediatric studies about the identification of SARS-CoV-2 through nasopharyngeal aspirate (NPA), but the use of NPA seems to be better than nasal or nasopharyngeal swab (NS) to identify respiratory virus in paediatrics.^16-19^ The NS is recommended because its collection is rapid and less traumatic, it is easy and more convenient to obtain, because it does not require training and additional devices; on the other hand, its sensitivity for the detection of respiratory viruses is lower compared to NPA.^16-19^ For this reason, we considered NPA as our reference test.

Our aim was to evaluate the concordance agreement and diagnostic performances of the NS compared to NPA, according to the age of the patients and the order in which tests were administered, for the purpose of evaluating if the NS could be used instead of NPA in the different conditions.

## METHODS

### Patients and samples

From March 13^th^ to May 22^nd^, all children who attended the emergency room and needed to be hospitalized and those who were transferred to our paediatric unit from other wards/hospitals, underwent NS and NPA performed from both nostrils, on admission and after 24 hours. The specimens were obtained by well-trained nurses or doctors. The nasopharyngeal swabs were collected following the procedure published on New England Journal of Medicine^20^, first from one nostril and then on the other side, using Copan-503CS01 nasopharyngeal flocked swab. The nasopharyngeal aspirates were collected from both the nostrils by using a standard protocol and a Medicoplast mucus extractor 440-ch08. There was not a defined order of obtaining the specimens. Particularly, we performed first the NS and then the NPA in March 2020; whereas we collected first the NPA and then the NS during the follow up.

A total of 134 patients were included. Thirteen among the latter and two outpatient children were followed collecting their paired specimens until both resulted negative 24 hours apart. Thus 300 paired specimens (NS/NPA) were collected from 136 patients (134 hospitalized and 2 outpatients) and were tested for SARS-CoV-2.

SARS-CoV-2 RNA was extracted using the Virus Kit Arrow Diagnostics – Italy and Elitech Diagnostics - Italy from the paired samples through nucleic acid amplification, performing the quantitative reverse transcription polymerase chain reaction (RT-PCR). The laboratory considered positive the samples in which were identified all the following genes: RNA-*polimerase, gene N (nucleocapside) and gene E (envelope); while the weak positivity was related to isolating one or two of the genes reported above*.

### Statistical analyses

All the statistical analyses were performed with R (v. 3.6.2)^21^. In order to estimate the incidence of the positive cases on hospitalized patients, we calculated the proportion of the positive cases (patients who had a positive result of NS or a positive result of NPA) in hospitalized patients and its 95% confidence interval using the binomial distribution.^22^

Analyses on diagnostic tests results (NS and NPA) concerned the concordance between the results of the two tests. Furthermore, we calculated the mismatch for positive and negative values and the sensitivity and specificity of NS (considering NPA as reference). In order to evaluate whether sensitivity, specificity and mismatch of the NS respect to the NPA were influenced by the age of patients, we considered the age as covariate (coded as 0 from 0 to 5 years old and as 1 for more than 5 years old).

Since the order of collecting specimens was different between patients hospitalized in March, and patient of the follow up, for the purpose of detecting any change in sensitivity, specificity and mismatches according to the order of execution of the tests, we considered the period as covariate (codified as 0 for the March period and as 1 for the Follow up).

Concerning the performance of NS in relation to NPA, descriptive summary measures were also calculated: likelihood ratio positive (LR+) and negative (LR-). Likelihood ratios compare the probability that a patient with NPA positive at the date of the test has a particular NS test result as compared to someone with NPA negative. LR+ is the ratio between the probability of a true positive result on the probability of false positive result. LR− is the ratio between the probability of a false positive result on the probability of a true negative result.^23^

Tests with very high LR+ and very low LR- have greater discriminating ability: in particular tests with LRs >10 or <0.1 are very useful in establishing or excluding a diagnosis.^24^

The analyses were performed by generalized estimating equation models (GEE) with family binomial to take account of the correlation among diagnostic tests on the same patients.^25^ To estimate the percentage of concordance, the model response was coded as 1 if the results of the two diagnostic tests agreed and as 0 otherwise. To estimate the sensibility and the mismatch for positive values only the records with a positive result of the NPA were used, and to estimate specificity and negative mismatch only the records with a negative result of the NPA were used. In both cases, the model response was the result of the NS (coded as 0 if it was negative and as 1 if it was positive).^26^

The influence of age and test period were tested by Wald test on the respective model coefficients with 5% significance level (two tailed test).

Due to the absence of a reliable value of prevalence of COVID-19 in children, we could not calculate the positive and negative predictive values (VPP, VPN).

## RESULTS

For clinical aims, we considered positive to SARS-CoV-2 every patient whose NPA or NS or NPA/NS resulted positive or weak positive.

Out of the 134 patients hospitalized, 18 children tested positive (prevalence 13.4%, 95%CI: 8.2%-20.4%); among the latter, 13 plus 2 outpatient children were followed collecting their paired specimens until both resulted negative 24 hours apart.

We collected 600 samples totally (300 paired): 43 positive NPA, 31 positive NS, 257 negative NPA and 269 negative NS.

Of the 300 paired specimens evaluated the naïve concordance was 92.0% (95% C.I.: 88.3%-94.6%).

The mismatch for negative results of NS for positive results of NPA was greater than the mismatch for positive values of NS for negative results of NPA (about 42% and about 2% respectively), see Table 1.

**Table 1.**
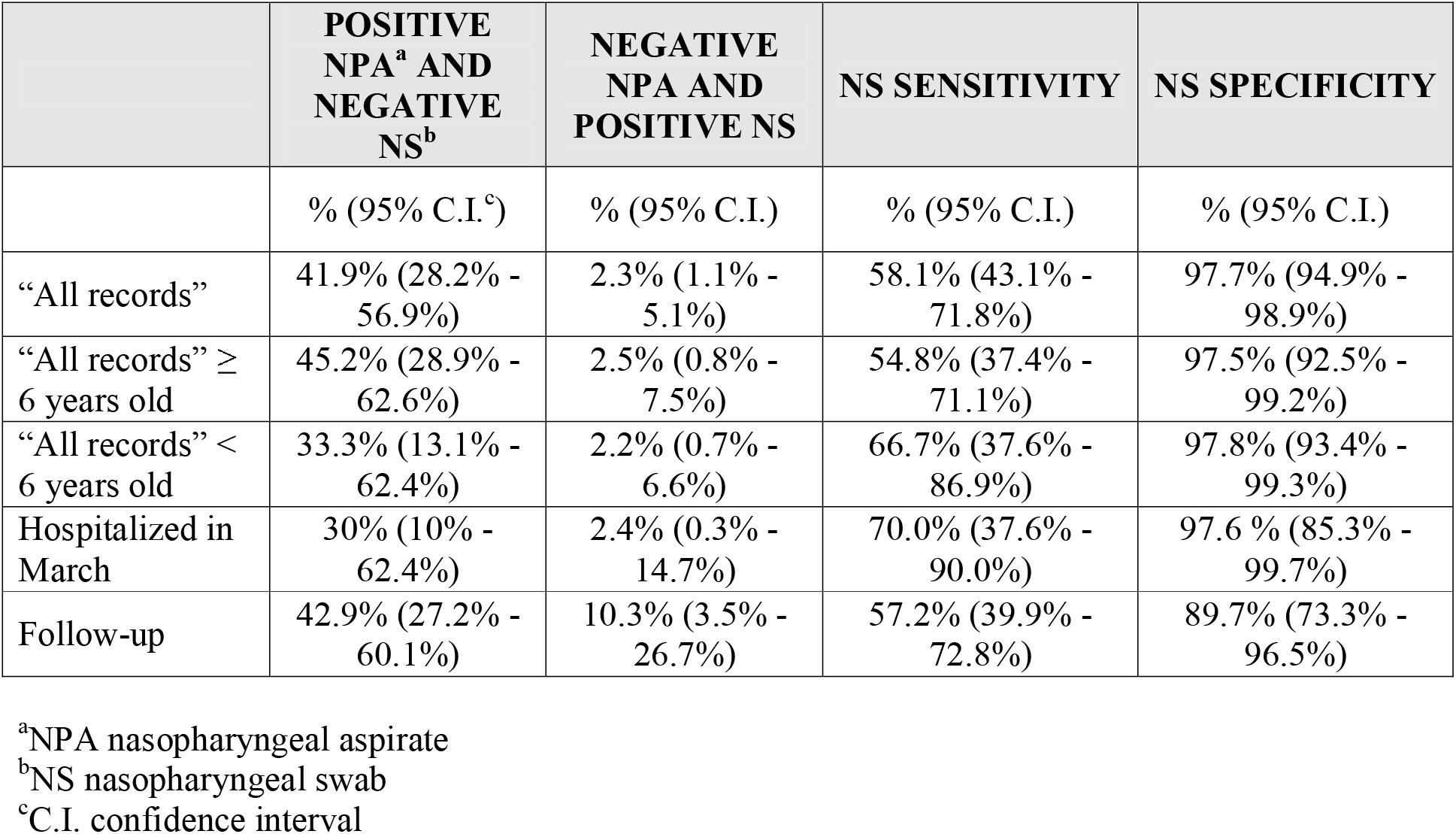
Sensitivity, specificity and mismatch for positive and negative results of Nasopharyngeal swab (NS) referred to Nasopharyngeal aspirate (NPA) in children.

The specificity of NS was greater than the sensitivity, suggesting the NS test was more suitable to rule-in positive NPA patients than rule-out negative NPA patients (sensitivity was about 58% and specificity was about 98%). The LR+ was 25.3 and LR- was 0.43.

The influence of age on the above-mentioned measures was not statistically significant: p=0.483 and 0.870 for positive and negative mismatch, and p=0.480, 0.870 for sensitivity and specificity.

The estimates of sensitivity, specificity, for the records of subjects ≥ 6 years old were similar to those for “all records” (Table 1); LR+ was 21.9 and LR- was 0.46. Considering the records for subjects < 6 years old, the mismatch for negative values of NS for positive results NPA (about 33%) was less than the results for the same analysis for all subjects and for subjects ≥ 6 years old; for the mismatch values of NS for negative results NPA (about 2%) we had similar results for all subjects and for subjects ≥ 6 years old (Table 1). The specificity of NS was greater than the sensitivity, suggesting the test was more suitable to detect positive NPA patients than negative NPA patients (sensitivity was about 67% and specificity was about 98%). In addition, sensitivity was greater than sensitivity calculated for all subjects and for subjects ≥ 6 years old (Table 1). The LR+ was 30.3 and LR- was 0.34.

As described above, we considered patients hospitalized in March (31 patients and 52 records) and patients involved in follow up (16 patients and 57 records).

Regarding hospitalized children in March, there were 52 paired specimens: 10 positive NPA, 8 positive NS, 42 negative NPA, 44 negative NS. Of the 52 paired specimens evaluated the naïve concordance was 92.3% (95% CI: 81.7% −97.0%). Results for these records showed the less mismatch for negative values of NS for positive results NPA (30%); but the mismatch for positive values of NS for negative results of NPA was similar to the results presented before (about 2%). Sensitivity was greater respect to the other results (70%), and the specificity was similar to the other results (about 98%), see Table 1. The LR+ was 29.2 and LR- was 0.31.

About the follow up samples, there were 57 paired specimens: 28 positive NPA, 19 positive NS, 29 negative NPA, 38 negative NS. Of the 57 paired specimens evaluated the naïve concordance was 73.7% (95%CI: 61.8%-82.9%). Results of mismatch for the negative values of NS and negative value of NPA showed similar results to the analyses about all subjects and for subjects ≥ 6 years old (about 43%); but the mismatch for positive values of NS for negative results of NPA was greater than all other results (about 10%). The specificity (about 90%) was greater than sensitivity (about 57%) and the specificity was the least of all the analyses (Table 1). The LR+ was 5.6 and LR-was 0.48.

The impact of period on the above-mentioned measures was not statistically significant for all models, in particular for the positive and negative mismatch we had p=0.471 and 0.180, and for sensitivity and specificity we had p=0.470 and 0.180 respectively.

## DISCUSSION

According to our results, the NS has a low sensitivity in the detection of SARS-CoV-2 in children. At the same time, NS has both high specificity and LR+; this means that positive NS has a good reliability in detecting who has positive NPA.

Although the effect of age and order of collection resulted not statistically significant, the difference observed between the results in the two age classes and in the two period of collection could suggested a potential impact of the two factors to be evaluated in larger case series.

Regarding the order of obtaining the tests, the sensitivity and specificity in hospitalized children in March were higher than those of the follow up patients, so with the collection of the NS before the NPA, there would be a major probability to identify SARS-CoV-2. One of the possible explanations is that NPA collects secretions from the lower parts of the upper respiratory tract compared to NS. Although both NPA and NS reach the nasopharyngeal tract, the thin catheter of the mucus extractor collect a larger number of deeper cells as demonstrated in different studies, in comparison with NS.^27,28^ Moreover, obtaining secretions firstly by suctioning with NPA, the remaining samples collected with NS might be reduced or inadequate to identify the virus. Nevertheless, our results were not statistically significant, because the 95% confidence interval was wide, probably due to the small size and not uniform sample examined. Further studies with a larger samples are necessary to strengthen this evidence.

Concerning the age, in the group of children younger than 6 years, the sensitivity was the highest of all the analysis. It means that negative NS has a good reliability in detecting the patient who has negative NPA. Moreover, patients younger than 6 years have the highest LR+ while they have the same specificity of all analyzed groups; so the NS is more suitable to find children with positive NPA. Performing NS in this group of age would be better for diagnosis of SARS-CoV-2.

However, in our experience, but in contrast with literature,^19,28^ performing NPA in young children, was simpler than NS: the aspiration of mucus with the small catheter into the nasopharynx resulted less unpleasant compared to the brushing against the nasopharyngeal wall caused by the NS. At the same time, NPA requires a catheter, an aspiration trap, a vacuum source and a specialized training, available only in a hospital setting. Instead, for NS no additional devices are needed.

Our study has some limitations. Firstly, the different order in obtaining the specimens; so the data are not uniform. Secondly, the small number of the sample. Another limitation is the lack of data about signs and symptoms of patients who underwent NPA and NS, and the resulting impossibility to describe a correlation between the isolation of SARS-CoV-2 and the clinical features. Finally, an analysis about the relationship between the viral load and the infectivity is not reported. It has been demonstrated that the identification of the virus on a specimen does not necessary correlate with infectivity, there are indeed multiple reports which attest a prolonged viral shedding after symptoms resolution in COVID-19.^29^ In our department, among children whose tests were positive, three continued to be positive to NS or NPA or both, for 9 weeks. These findings agree with several studies which demonstrated the prolonged viral shedding of children^30,31^.

## CONCLUSIONS

The NS has a low sensitivity in detecting SARS-CoV-2 in children when referred to NPA, both in the overall analysis and in that according to the age; instead its specificity results high. It means that a positive NS can be reliable, but that a negative NS cannot rule out the presence of SARS-CoV-2 since the proportion of false negatives is substantial.

Though statistically not significant, the sensitivity of NS becomes higher if it is performed before NPA, as the patients hospitalized in March were undertaken.

Our results suggest to prefer the collection of NS whenever possible for the detection of SARS-CoV-2 in children younger than 6 years, thanks to its high specificity, even though we did not reach the statistical significance.

As far as we know, this is the first study dealing with comparing NS to NPA for detecting SARS-CoV-2 and further analysis are mandatory to transfer these findings to our clinical practice.

## Data Availability

All data referred to in the manuscript are available

## Acknowledgments

This research received no specific grant from any funding agency in the public, commercial, or not-for-profit sectors.

## Authors’ contributions

Prof Marchisio, Castaldi and Biganzoli conceptualized and designed the study, reviewed and revised the manuscript. Drs Di Pietro and Capecchi collected data, wrote and reviewed the manuscript. Drs Biganzoli, Luconi, Marano and Boracchi analyzed the statistical data. Drs Lunghi and Ceriotti analyzed the specimens. Prof Gori and Agostoni, Drs Bertolozzi, Bosis, Pinzani, Cantoni reviewed the manuscript. All authors approved the final manuscript as submitted and agree to be accountable for all aspects of the work.

